# A method to reduce ELISA serial dilution assay workload applied to SARS-CoV-2 and seasonal HCoVs

**DOI:** 10.1101/2021.09.13.21263523

**Authors:** David Pattinson, Peter Jester, Lizheng Guan, Seiya Yamayoshi, Shiho Chiba, Robert Presler, Hongyu Rao, Kiyoko Iwatsuki-Horimoto, Nobuhiro Ikeda, Masao Hagihara, Tomoyuki Uchida, Keiko Mitamura, Peter Halfmann, Gabriele Neumann, Yoshihiro Kawaoka

## Abstract

**Objectives:** Assays using ELISA measurements on serially diluted serum samples have been heavily used to measure serum reactivity to SARS-CoV-2 antigens and are widely used in virology and elsewhere in biology. We test a method to reduce the workload of these assays, and measure reactivity of SARS-CoV-2 and HCoV antigens to human serum samples collected before and during the COVID-19 pandemic.

**Methods:** We apply Bayesian hierarchical modelling to ELISA measurements of human serum samples against SARS-CoV-2 and HCoV antigens.

**Results:** Inflection titers for SARS-CoV-2 full-length spike protein (S1S2), spike protein receptor-binding domain (RBD), and nucleoprotein (N) inferred from three spread-out dilutions correlated with those inferred from eight consecutive dilutions with an R^2^ value of 0.97 or higher. We confirm existing findings showing a small proportion of pre-pandemic human serum samples contain cross-reactive antibodies to SARS-CoV-2 S1S2 and N, and that SARS-CoV-2 infection increases serum reactivity to the beta-HCoVs OC43 and HKU1 S1S2.

**Conclusions:** In serial dilution assays, large savings in resources and/or increases in throughput can be achieved by reducing the number of dilutions measured and using Bayesian hierarchical modelling to infer inflection or endpoint titers. We have released software for conducting these types of analysis.

## Introduction

Severe acute respiratory syndrome coronavirus 2 (SARS-CoV-2) emerged in December 2019 [1] and is the causative agent of coronavirus disease 2019 (COVID-19). On March 11, 2020, the outbreak was declared a pandemic by the World Health Organization (WHO) [2] and at the time of writing there have been over 206 million confirmed SARS-CoV-2 infections and 4.3 million deaths attributed to the virus globally [3]. The World Bank estimates that between 119 and 124 million people were pushed into extreme poverty by impacts of the COVID-19 pandemic in 2020 alone [4].

Coronaviruses (CoVs) more generally are separated into four phylogenetic clades: alpha-CoV, beta-CoV, gamma-CoV and delta-CoV. Four CoVs circulate continuously in humans (HCoVs), are distributed globally, and generally cause common cold or influenza-like illness [5]. These are the alpha-CoVs HCoV-229E (229E) and HCoV-NL63 (NL63), and the beta-CoVs HCoV-HKU1 (HKU1) and HCoV-OC43 (OC43). Two other beta-CoVs have also infected humans: severe acute respiratory syndrome coronavirus (SARS-CoV) and Middle East Respiratory Syndrome Coronavirus (MERS-CoV) [6,7]. The SARS-CoV outbreak in 2003 was largely contained and MERS-CoV is not well adapted for ongoing human to human transmission.

At the start of the COVID-19 pandemic we lacked detailed knowledge of serological reactivity to HCoVs in the human population. It was also unknown how much cross-reactivity there was between antibodies elicited to prior HCoV infections against SARS-CoV-2. Furthermore, if cross-reactivity did exist, it was not known how protective it may be against SARS-CoV-2 infection. Several studies have now measured reactivity of human serum collected before and during the COVID-19 pandemic, and through the course of SARS-CoV-2 infections against panels of HCoV antigens [8–17]. Most studies that measure human sera collected before the COVID-19 pandemic demonstrate that a small proportion of pre-pandemic sera possess cross-reactive antibodies against the SARS-CoV-2 S1S2 and N antigens [8,9,15,17]. Moreover, studies that measure sera collected longitudinally from COVID-19 patients show that SARS-CoV-2 infection increases serum antibody titers to certain beta-HCoV antigens, particularly OC43 [10–12,14]. This observation tends to be weaker, absent, or even reversed when comparing cohorts of unmatched samples collected before and during the pandemic [9,15,18]. The diverse nature of cohorts and variable circulation of HCoVs when cohorts were sampled may contribute to the sometimes conflicting reports [19].

Serology studies can be conducted using a single serum dilution in ELISA assays. This enables high sample throughput but can limit resolution if the dilution used results in measurements near to the minimum or maximum Optical Density (OD) readout of the assay. Thus, it is common to measure the OD across a series of typically 8-12 dilutions and report the dilution, known as the ‘endpoint’, at which the OD drops below a predefined baseline value. However, conducting ELISAs at this many serial dilutions per sample is labor- and resource-intensive. Here, we investigated if equally accurate data can be obtained from substantially fewer than 8-12 serial dilutions, by using a Bayesian hierarchical model to fit sigmoid curves to the data.

Sigmoid curves are used to fit OD values as a response to log serum concentration (Fig. S1) [20]. Once the model is fitted, quantities of interest, such as the inflection point, or any arbitrary endpoint, can be derived from the parameters describing the curve. This approach offers several benefits. Firstly, sigmoid curves can be inferred from fewer dilutions than are typically conducted. Secondly, fitting a model to multiple measurements reduces the influence of error associated with individual measurements, avoiding reliance on individual measurements used to define “the” endpoint. Thirdly, parameters are estimated on a continuous scale in between discrete dilution folds, increasing resolution. Fourthly, samples from ELISA datasets often share properties such as the maximal response (‘saturation’ point of the ELISA), minimal response (the OD value of the background or negative control) and gradient of the sigmoid curve. When properties are shared among samples, hierarchical modelling allows measurements from all samples to contribute to the inference of the shared properties. Data from a single sample does not have to be able to accurately infer these properties in isolation; only parameters of interest, such as the horizontal shift of the sigmoid, need to be inferred on a per-sample basis.

As stated earlier, most ELISA studies report an ‘endpoint’ titer, defined as the first dilution in the series at which an OD value below a cutoff is measured. Cutoffs are determined by either choosing a low absolute OD value, computing a low proportion of the maximal response, or computing a multiple of the mean background or negative control. Frey et al. [21] advocate a less arbitrary method based on computing the prediction limit of the negative control. The primary variation of interest when comparing dilution series between samples is the extent to which the sigmoid curves are shifted to the left or right on the x-axis (Fig. S1). The location of a sigmoid curve relative to the x-axis is defined by its inflection point (parameter a, Fig. S1). In the context of a titration assay we refer to this inflection point as the ‘inflection titer’. Rather than comparing arbitrarily defined endpoint titers, we focus our analysis primarily on comparing inflection titers. We note that once the parameters of a sigmoid curve are inferred, it is elementary to compute the endpoint at any arbitrary cutoff value.

Here, we tested serum samples from several cohorts (collected before the SARS-CoV-2 pandemic, collected from individuals who tested PCR-positive for SARS-CoV-2, and collected from individuals early and late during SARS-CoV-2 infection) in ELISAs against SARS-CoV-2 and seasonal HCoVs. This comprehensive dataset was then used to test if a reduced number of serial dilutions can be used to accurately infer inflection titers in a Bayesian hierarchical modelling framework. We demonstrate that only three serum dilutions are sufficient to recapitulate inflection titers obtained with eight serum dilutions, resulting in substantial time and resource savings.

## Materials Methods

### Human serum samples

248 samples were purchased commercially before the emergence of SARS-CoV-2 (64 from Lampire Biological Laboratories, 184 from BioIVT). 188 samples were collected from individuals at the University of Wisconsin-Madison Hospital (UWH) after testing PCR-positive for SARS-CoV-2; no further tests (such as serology test or virus isolation) were conducted to confirm the PCR results. 12 SARS-CoV-2 positive samples were purchased from Lampire Biological Laboratories (commercial positive). 44 paired samples were collected at the Eiju General Hospital (EGH), Tokyo, Japan early and late during SARS-CoV-2 infection. Early and late samples were collected on average 2.0 (standard deviation 4.8) and 29.5 (standard deviation 9.6) days after symptom onset, respectively. Four samples were from asymptomatic patients, for which the day of RT-qPCR positive test was used as the day of onset. 162 samples were collected by the Marshfield Clinic Research Institute (MCRI), Marshfield, Wisconsin after the emergence of SARS-CoV-2. Groups of samples used in this study are summarized in Table S1.

### Ethics and biosafety statements

University of Wisconsin: The study was approved by the Human Subjects Institutional Review Boards at the University of Wisconsin and signed informed consent was obtained from each participant at each time point before collection. MCRI: Human samples were collected after informed consent by protocols approved by the MCRI’s Institutional Review Board. IMSUT: Human samples were collected by following protocols approved by the Research Ethics Review Committee of the Institute of Medical Science, the University of Tokyo (approval number 2019 - 71-0201). Signed informed consent was obtained from all participants.

### Antigens

SARS-CoV-2 full-length spike protein (S1S2) and its receptor-binding domain (RBD) were expressed in Expi293F cells and purified using TALON metal affinity resin (Thermo Fisher Scientific) following the Krammer group protocol [22]. The S1S2 construct included the ‘hexaproline’ stabilizing amino acid (AA) substitutions (F817P/A892P/A899P/A942P/K986P/V987P) [23], the cleavage site RRAR (682-685) was replaced with GSAS, and AA15-1208 was followed by T4 foldon trimerization motif, 3C protease cleavage site, and hexa-histidine tag (AA1-14 is a signal peptide). AA319-541 were used as the RBD. Other antigens were purchased from Sino Biological (OC43 S1S2, catalog number 40607-V08B; HKU1 S1S2, 40606-V08B; NL63 N, 40641-V07E; 229E N 40640-V07E; SARS-CoV-2 N 40588-V08B), BPS Bioscience (NL63 S1S2, 100788), Creative Diagnostics (OC43 N, DAGB101), The Native Antigen Company (229E S1S2, REC31880-100) and Aviva Systems Biology (HKU1 N, OPCA203310).

### ELISA

#### IRI

ELISA on pre-pandemic, UWH PCR positive, commercial positive and MCRI samples was conducted at the Influenza Research Institute (IRI), University of Wisconsin-Madison, as follows. 96-well ELISA plates (Thermo Scientific #:14245153) were coated with antigen at 100 ng/well and incubated overnight at 4 °C. Plates were blocked with PBS containing 0.1% Tween-20 (PBS-T) and 3% milk for 1 h at room temperature and washed. All washes were conducted three times with PBS-T. Sera were diluted 1:40 in PBS-T and 1% milk. For titration series, sera were further diluted in PBS-T and 1% milk in 1:4 steps to a lowest concentration of 1:655,360. 100 µl of each sera was added to the ELISA plates and incubated for 2-4 h at room temperature. Anti-human IgG-Peroxidase antibody produced in goat (Sera Care-KPL #.5220-0277), diluted 1:3000 in PBS-T and 1% milk, was used as the secondary antibody. Plates were washed and incubated for 1 h at room temperature with 50 µl/well of diluted secondary antibody. Plates were washed, and then incubated with 100 µl OPD solution (SIGMAFAST™ OPD, Sigma-Aldrich #P9187) for 10 min at room temperature. The reaction was stopped by adding 50 µl 3 M HCl and the OD at 490 nm was measured.

#### IMSUT

ELISA on EGH samples was conducted at the IMSUT as described previously [24]. 96-well Maxisorp microplates (Nunc) were incubated with 100 ng/well of recombinant protein or PBS at 4°C overnight and then incubated with 5% skim milk in PBS containing 0.05% Tween-20 for 1 h at room temperature. Serum samples were diluted 1:40 in PBS-T containing 5% skim milk. Microplates were reacted for 1 h at room temperature with the diluted serum samples, followed by peroxidase-conjugated goat anti-human IgG, Fcγ Fragment specific antibody (Jackson ImmunoResearch #109-035-098). 1-Step Ultra TMB-Blotting Solution (Thermo fisher scientific) was then added to each well and incubated for 3 min at room temperature. The reaction was stopped by adding 2 M H_2_SO_4_ and the OD at 450 nm was measured.

### Statistical analyses

At least two blank wells (PBS only) were run on each 96-well plate during ELISA measurement. The mean of the blank values on a plate was subtracted from other values on a plate to correct for background. Bayesian inference was used to estimate posterior parameter distributions for all analyses. Eight chains each took 10,000 samples using the NUTS sampler implemented in pymc3 [25]. Excellent convergence was achieved in all analyses.

#### Sigmoid curves

A four parameter logistic curve was used to model the OD of sample i (y_i) as a response to log dilution (x_i) (Fig. S1) [26]: y_i ∼ dnorm(c + d_i / (1 + exp(-b(x_i - a_i)), s). x_i was calculated as log_4(t/40) where t is the dilution. a (the horizontal location of the sigmoid) was modelled with no pooling between samples whereas b (the gradient) and d (the difference between minimum and maximum response) were modelled with partial pooling between samples. Given that OD values were background adjusted, c (the minimum response) was fixed at zero. Graphical interpretations of each parameter is shown in Fig. S1. Weakly informative priors were used: a_i ∼ dnorm(u_a, s_a), b ∼ dnorm(0, 1), d_i ∼ dexp(s_d), u_a ∼ dnorm(0, 1), s_a ∼ dexp(1), s_d ∼ dexp(1). Log dilution was standardized to have mean of zero and standard deviation of one for all inference. Inflection titers are computed using the mean value of the posterior distribution of a for each sample as: 40(4 ** a_i).

#### Seroconversion probabilities

We used robust logistic regression to estimate the probability of seroconversion based on ELISA OD values measured against SARS-CoV-2 RBD and S1S2 antigens at a dilution of 1:40. Negative (pre-pandemic samples) or positive SARS-CoV-2 infection status (PCR-positive samples) was encoded as zero or one, respectively, and modelled as: y_i ∼ dbern(alpha / 2 + (1 - alpha) f(b_0 + b_R x_Ri + b_S x_Si)). Where b_R and b_S are the effect size estimates of RBS and S1S2 and x_Ri and x_Si is the OD value against RBD or S1S2 in individual i, f is the standard logistic function: f(x) = 1/(1+exp(-x)). The regression is ‘robust’ due to alpha; when alpha = 1, the probability a sample is positive is 0.5 and independent of the b coefficients or OD values. Outliers, such as those caused by false-positive PCR, are accounted for by this random component without impacting the fit of the logistic curve. Weakly informative priors were used: alpha ∼ dbeta(1, 4) based on the assumption that outliers should be rare; and beta_j ∼ dnorm(0, 1). OD values were standardized to have a mean of zero and standard deviation of 1. Samples were partitioned into training and test sets in a ratio of 1:4. Partitioning was stratified so that the ratio of negative to positive samples in the validation set was as close as possible to that in the training set.

#### Testing difference of group means

We used a linear model to test the difference of means between two groups. The response (y_i) was modelled as: y_i ∼ dstudentt(b_0 + b_1 x_i, nu, sigma). Weakly informative priors were used: b_0 ∼ dnorm(0, 1), b_1 ∼ dnorm(0, 1), nu ∼ dexp(1), sigma ∼ dexp(1). x_i is a binary array indicating group membership of sample i. Data were standardized to have unit variance and a mean of zero. The Student-T, rather than the Normal, distribution was used to be less sensitive to outliers.

#### Testing differences among groups of paired samples

We computed the difference between the early and late infection ELISA OD measurements for each individual, delta_i, by subtracting the early value from the late. We then inferred the parameters of a Student-T distribution to describe these differences: delta_i ∼ dstudentt(mu, nu, sigma). These priors were used: mu ∼ dnorm(0, 1), nu ∼ dexp(1), sigma ∼ dexp(1). The Student-T, rather than the Normal, distribution was used to be less sensitive to outliers.

## Results

We tested human serum samples collected before and during the SARS-CoV-2 pandemic for antibodies to SARS-CoV-2 and HCoVs, and used this well-characterized dataset to test a computational approach for substantially reducing the number of serial dilutions needed to robustly measure inflection points (and other arbitrary endpoints) of titration series.

### Reactivity of pre-pandemic and SARS-CoV-2 PCR-positive human sera with SARS-CoV-2 antigens

First, we measured the reactivity of human serum samples collected before the pandemic and from individuals who were PCR-positive for SARS-CoV-2 against SARS-CoV-2 S1S2, RBD and N antigens at a single serum dilution of 1:40 (Fig. 1A). Most of the PCR-positive individuals had relatively high OD values against all three SARS-CoV-2 antigens (Fig. 1A); however, a small minority did not react to SARS-CoV-2 antigens which may have been caused by false-positive PCR results. As expected, most pre-pandemic sera lacked reactivity to SARS-CoV-2 antigens (Fig. 1A). A robust logistic regression was fit to the 80% of the samples (the training set), and used to infer the seropositivity rate. In the test set, 95% of samples classified as positive (i.e., the assay sensitivity) and 98% classified as negative (i.e., the assay specificity) were correctly identified. Summarizing all pre-pandemic samples, 2.02% were SARS-CoV-2 RBD seropositive, 1.61% were SARS-CoV-2 S1S2 seropositive, and 12.9% were SARS-CoV-2 N seropositive, consistent with several published studies [8,9,15].

**Fig. 1.**
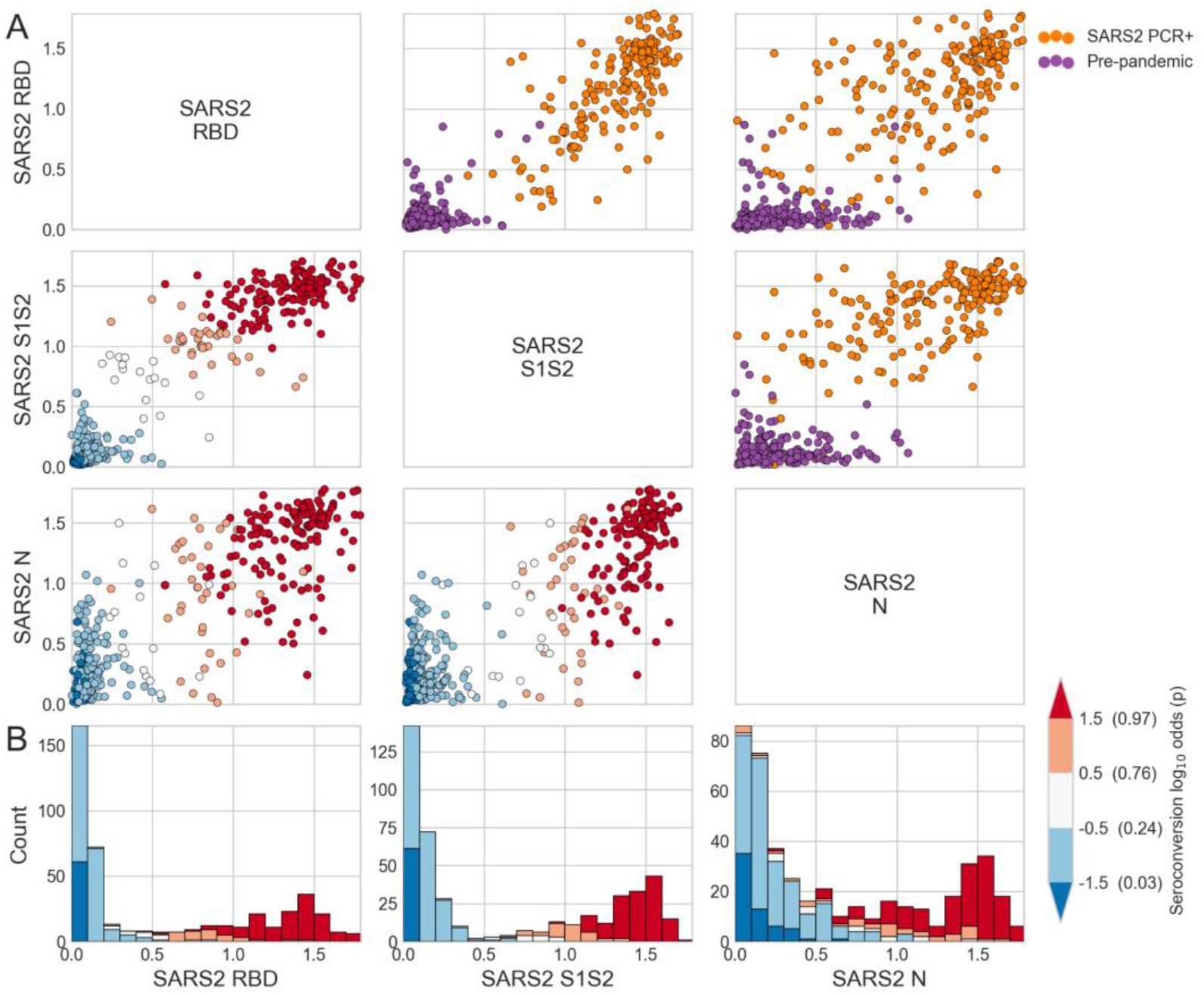
Reactivity to SARS-CoV-2 (SARS2) spike and N antigens in pre-pandemic and SARS-CoV-2 PCR-positive human serum samples. **(A)** Scatter matrix of ELISA OD values of human serum samples collected before the outbreak of SARS-CoV-2 and from individuals that tested PCR-positive for SARS-CoV-2 measured against three SARS-CoV-2 antigens. Individual columns and rows show OD values of one SARS-CoV-2 antigen (RBD, S or N). Subplots above the main diagonal are colored by binary infection status. Subplots below the main diagonal are colored by seroconversion log odds. **B**) Histograms display the distribution of measurements for individual antigens and are colored by seroconversion log odds.

### Reactivity of pre-pandemic and SARS-CoV-2 PCR-positive human sera with HCoV antigens

To measure the reactivity of unpaired pre-pandemic and SARS-CoV-2 PCR-positive samples to HCoVs, we conducted ELISAs at a single dilution of 1:40 against the S1S2 and N antigens of the two beta-HCoVs (OC43 and HKU1) and the two alpha-HCoVs (NL63 and 229E, Fig. 2). Conducting a single dilution, rather than multiple, allows higher throughput but may limit assay resolution by potentially yielding OD values that are close to the minimum or maximum OD for the assay. Most sera had high OD values to HCoVs, indicating that most individuals have been infected with all four HCoVs, consistent with earlier studies [27-30]. For these unpaired sets of samples, we found very minor or no differences between pre-pandemic and SARS-CoV-2 PCR-positive sera measured against the S1S2 of all HCoVs (Fig. 2). For the N protein, the beta-HCoVs OC43 and HKU1 showed lower reactivity to SARS-CoV-2 PCR-positive sera than pre-pandemic samples, and the alpha-HCoVs showed either no difference (NL63) or a small increase (229E) (Fig. S2).

**Fig. 2.**
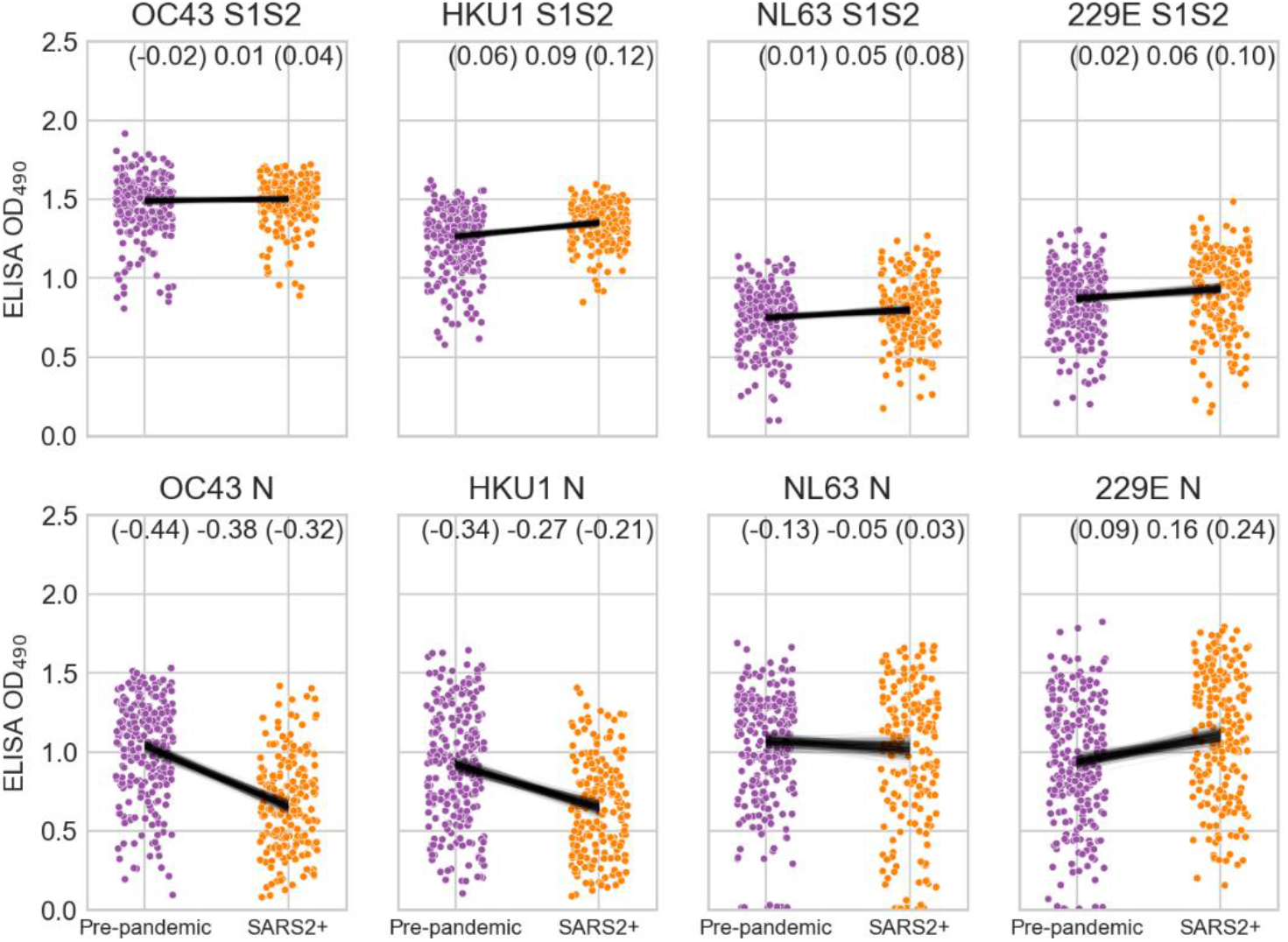
Reactivity of pre-pandemic and SARS-CoV-2 PCR-positive human sera (SARS2+) to HCoV antigens. ELISA OD values for 1:40 diluted serum against HCoV S1S2 and N proteins. Black lines are the posterior distribution of differences in group means inferred from a linear model using Student-T distributed error. The upper and lower boundaries and mean of the 95% highest posterior density interval of difference in group means is shown in the top right.

We also investigated the impact of SARS-CoV-2 infection on OC43 and HKU1 S1S2 and N antigen reactivity using paired human serum samples from COVID-19 patients collected early and late (mean 2.0 and 29.5 days after symptom onset respectively) during SARS-CoV-2 infection (Fig. 3). OD values against OC43 and HKU1 S1S2 proteins increased between early and late samples, demonstrating that SARS-CoV-2 infection increases reactivity to the full-length spike proteins of beta-HCoVs, consistent with other studies [27-30]. There was no systematic increase in OD values for the HKU1 S1 protein (lacking the S2 domain) in ELISA measurements from early to late samples, indicating that the increase in OD values measured against the full-length spike is caused by the S2 domain (Fig. 3).

**Fig. 3.**
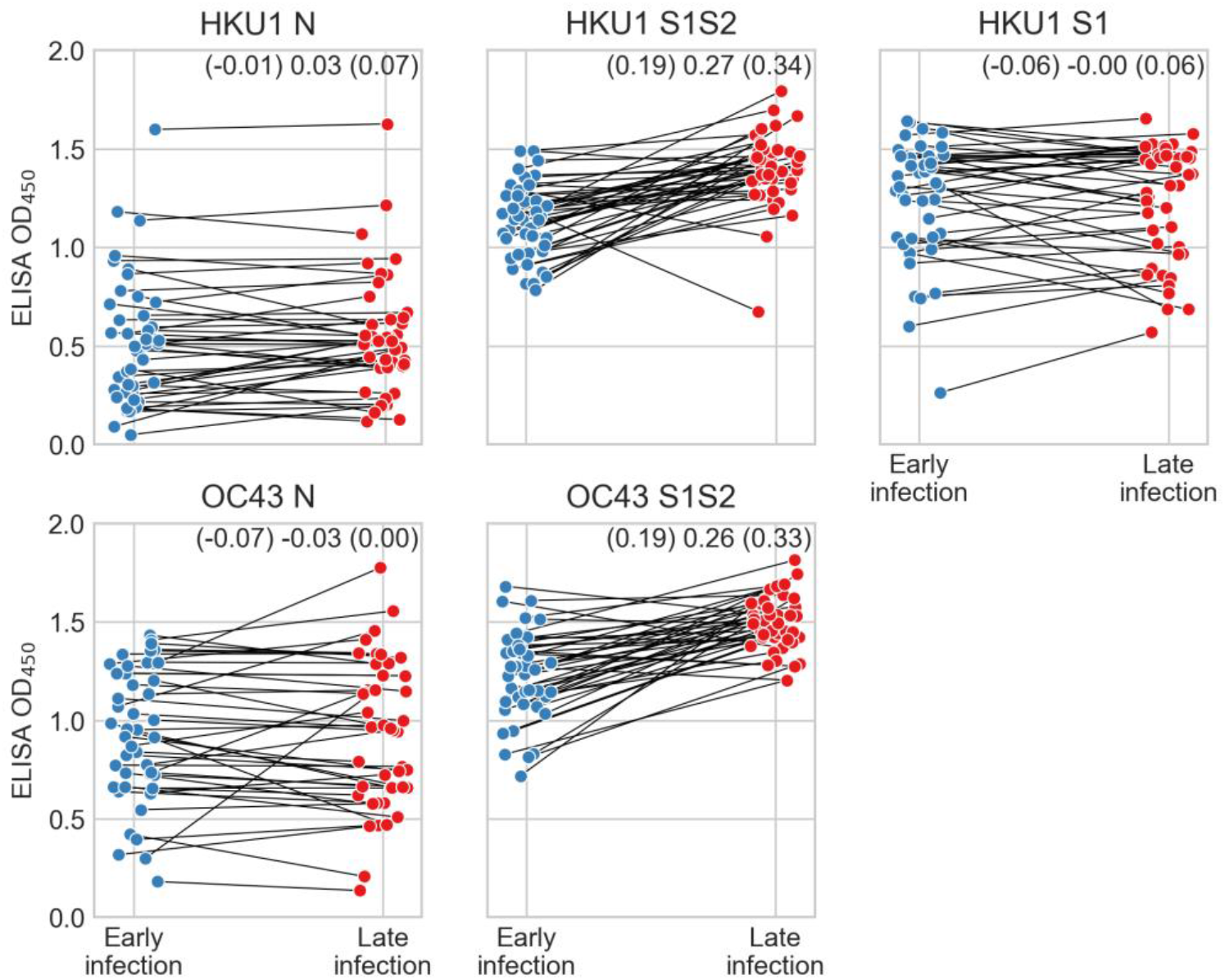
Reactivity to beta HCoV antigens during SARS-CoV-2 infection. ELISA reactivity of human antisera collected early and late during SARS-CoV-2 infection against beta-HCoV proteins. Lines connect early and late samples from the same individuals. Student-T distributions were fit to paired differences between late and early infection measurements. Upper and lower boundaries and mean of the 95% highest posterior density interval of the Student-T distribution mean are indicated.

### Serum reactivity analysis with a limited number of serum dilutions

As stated previously, multiple ELISA measurements from serial dilutions of serum are more informative than measurements based on a single serum dilution but are labor- and resource-consuming. Here, we investigated if less than eight dilutions can be used to robustly infer inflection and endpoint titers.

For this task, we used 162 human sera with unknown SARS-CoV-2 exposure status collected by the MCRI during the pandemic. First, ELISAs were conducted at eight four-fold dilutions starting at 1:40 against SARS-CoV-2 S1S2, RBD, and N. Then, we inferred parameters of sigmoid curves for each sample using all eight dilutions in a Bayesian hierarchical model, and derived inflection titers. Finally, we tested if similar inflection titers can be obtained using subsets of the eight dilutions. Inflection titers can be calculated with high confidence from just three ‘spread-out’ dilutions (1:40, 1:640, 1:10240; Fig. 4). Highest posterior density intervals (HPDIs) of inflection titers increased at higher titers when only two dilutions (e.g., 1:40 and 1:640) or three consecutive dilutions (e.g., 1:40, 1:160, 1:640) were used (Fig. 4). However, including one additional dilution (1;40, 1:160, 1:640, 1:2560) or spreading out dilutions (1:40, 1:640, 1:10240) reduces HPDI width, even at high inflection titers (Fig. 4). Thus, highly robust analyses can be conducted from as few as three serum dilutions. To further evaluate our approach, we compared an arbitrarily chosen endpoint cutoff (OD=0.1) calculated from subsets of three or four dilutions to eight dilutions (Fig. S3). As with inflection titers, endpoint titers inferred from only three serum dilutions correlated very closely with those inferred from eight serum dilutions (R^2^≥0.97).

**Fig. 4.**
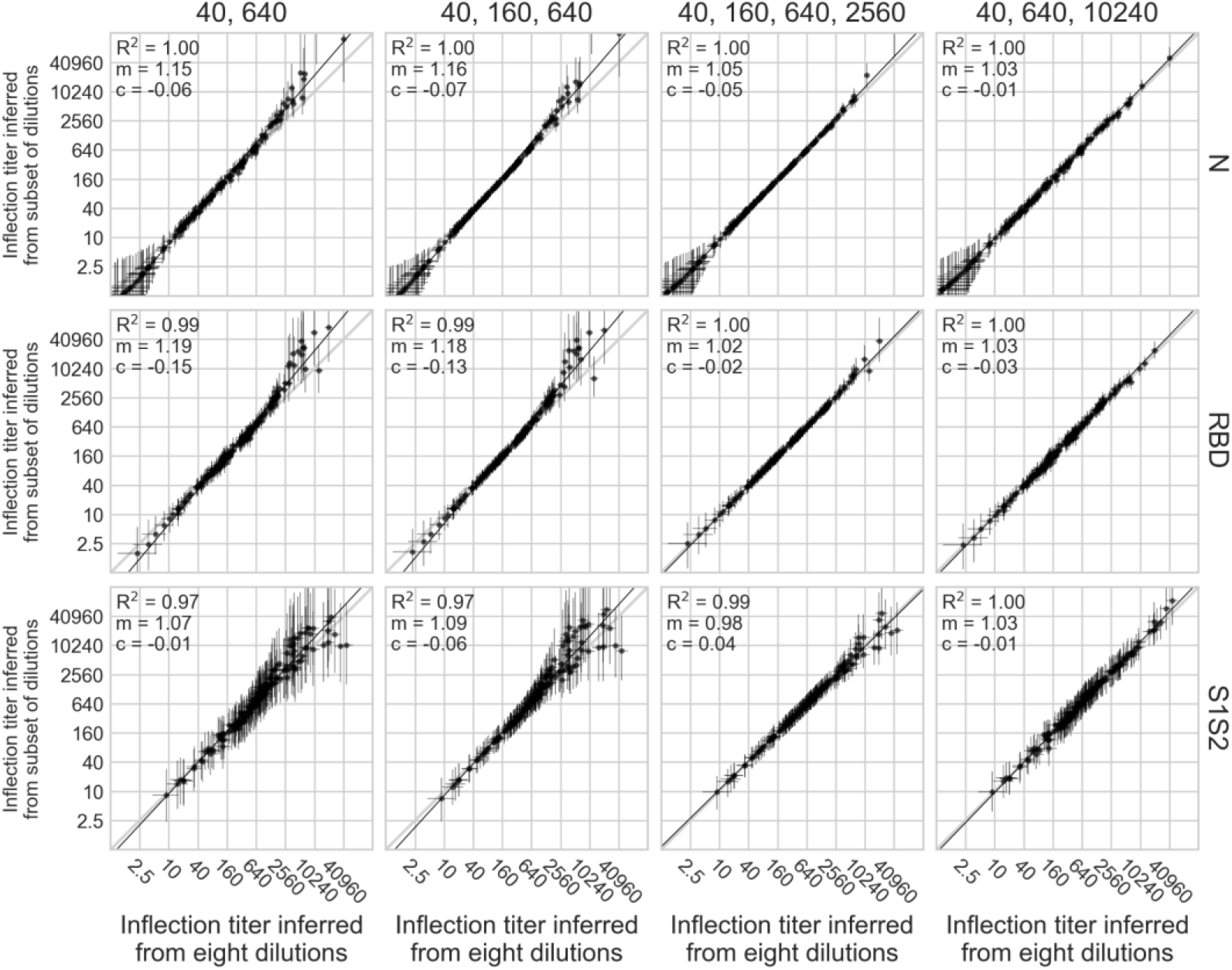
Inferring inflection titers from minimal dilution series. Inflection titers were computed using eight four-fold dilution series starting at 1:40; this value is shown on the x-axes. Inflection titers were also computed using different subsets of dilutions: (40, 640), (40, 160, 640), (40, 160, 640, 2560) and (40, 640, 10240); these values are shown on the y-axes. Ordinary least squares linear regression was conducted on each x and y series. R squared, slope (m) and intercept (c) are indicated on each plot. Error bars indicate the 95% highest posterior density interval of the posterior distribution of the inflection titer.

To test our approach further, we measured ELISA OD values using a dilution series of 1:40, 1:640, and 1:10240 to infer inflection titers for the S1S2 protein of the beta-HCoVs HKU1 and OC43 against pre-pandemic and SARS-CoV-2 PCR-positive human serum samples. Inflection titers were higher in SARS-CoV-2 PCR-positive samples than pre-pandemic samples (Fig. 5). This is different to the pattern shown in Fig. 2 where there is no difference between pre-pandemic and PCR-positive samples in OC43 S1S2 and only a very small difference in HKU1 S1S2. This illustrates the importance of conducting measurements across serial dilutions, especially if the single dilution used generates OD values close to the minimum or maximum OD value of an assay. In this case, at a dilution of 1:40 the OD readout is close to saturation which obscures the differences between groups (Fig. S4).

**Fig. 5.**
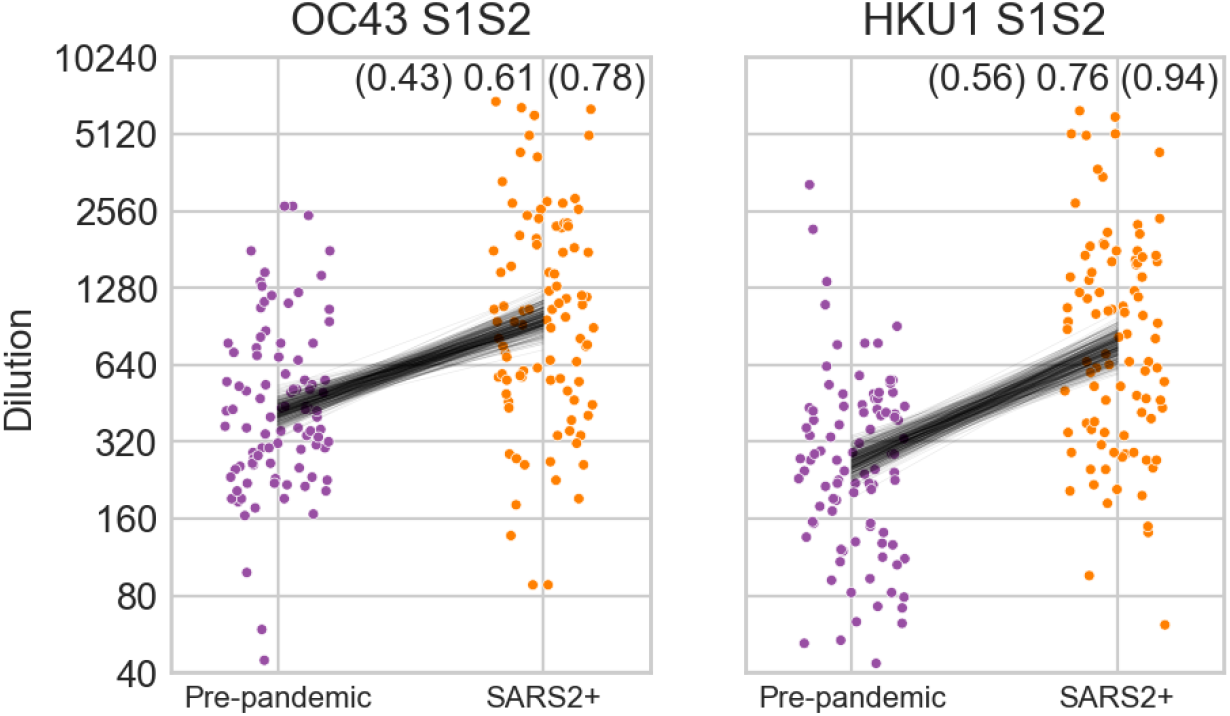
Inflection titers of pre-pandemic and SARS-CoV-2 PCR positive human sera (SARS2+) to beta-HCoV S1S2 proteins. Black lines show the posterior distribution of differences in group means inferred from a linear model using Student-T distributed error. The upper and lower boundaries and mean of the 95% highest posterior density interval of difference in group means is shown in the top right; one unit is a single four-fold dilution.

## Discussion

Here, we used human sera collected during the SARS-CoV-2 pandemic to demonstrate that it is possible to robustly infer inflection and endpoint titers from fewer than the typically used 8-12 serial dilutions. In fact, three widely spaced dilutions (1:40, 1:640, 1:10240) provide the same information as eight consecutive dilutions. Thus, researchers could conduct a pilot study on a small number of samples with 8-12 dilutions, infer inflection (or endpoint) titers, and identify a subset of dilutions that can be used to obtain equally accurate results. Routine analysis with large sets of sera could then be carried out with only the minimal set of dilutions.

Our approach is based on fitting sigmoid curves, rather than simply identifying the serum dilution at which a predetermined endpoint is reached. This approach has several advantages; we show it substantially reduces the number of dilutions required, reducing resources, effort and/or increasing throughput. It also generates readings on a continuous scale between discrete dilution folds, increasing assay resolution. Furthermore, multiple data points are used to fit sigmoid curves, meaning individual readings with large error have a smaller impact on the final readout. In systems prone to large sources of error, specific processes for accounting for the error can be included [31].

Hierarchical modelling that enables shared inference of parameters between samples imparts additional benefits and is a large factor in being able to reduce the number of dilutions required. For instance, if all samples tend to the same maximal response at high concentration, as is the case in this study, it is unnecessary to ensure that the dilution series allows the same maximal response to be inferred in every sample independently. Rather, dilutions should be positioned in the variable region of the response, and inference of the maximal response should be fully pooled. Additional modelling choices can further reduce dilutions required. In this system it was known *a priori* that OD values tend to zero as concentration decreases (after subtracting the background reading). Therefore, it is unnecessary to conduct measurements at very low concentration because this information can be expressed in the model by setting c=0.

Reducing the number of dilutions required to infer parameters of a sigmoid curve is not restricted to analysis of ELISA OD measurements. Assays conducted on a serially diluted sample are common in virology [32,33] and widespread in biology generally [20]. When applying this approach to novel systems, choice of which parameters are fully versus partially pooled, or fixed at a certain value, depends on the nature of the assay, the data, and the question being addressed. Pilot experiments should guide choice of a dilution series that will allow accurate inference (Fig. 4).

We have released R and python packages to fit sigmoid curves using minimal datasets (https://github.com/IRI-UW-Bioinformatics/inflection-titer). Other dose-response curve packages exist [20], however this implementation specifically focuses on hierarchical modelling across samples using Bayesian inference [25,34], both features which are absent from existing packages [20].

Here, we measured cross-reactivity to SARS-CoV-2 S1S2, RBD, and N antigens in human serum samples collected before the emergence of SARS-CoV-2 and found similar rates as those reported elsewhere [8,9,15]. We also measured how patterns of reactivity to seasonal HCoVs changed as a result of the SARS-CoV-2 pandemic. We see robust increases in reactivity to seasonal beta-HCoVs in two different comparisons: pre-pandemic versus SARS-CoV-2 PCR-positive sera (Fig. 5), and sera collected early versus late after a confirmed SARS-CoV-2 infection (Fig. 3). The latter demonstrates that the increase in beta-HCoVs antigen titers is the result of SARS-CoV-2 infection, rather than individuals with existing high titers to beta-HCoVs being more likely to be infected with SARS-CoV-2. This effect is masked in our analysis of 1:40 diluted sera (Fig. 2) because most measurements are near the saturation OD reading (Fig. S4). Our findings are consistent with other reports, although some differences exist among the studies: while several studies reported increases in HKU1 and OC43 spike protein titers in pandemic compared to pre-pandemics samples [9–11], others found increased OC43 (but not HKU1) reactivity [16]. Such differences may be a consequence of the diverse nature of cohorts used and variable circulation of individual HCoVs during the years that samples were collected.

## Data Availability

Data is available on request.

## Acknowledgements

The authors would like to thank Huong McLean, Jennifer King, Ed Belongia of the MCRI, Marshfield, Wisconsin, USA, for human serum sample collection.

## Supplementary Material

**Fig. S1.**
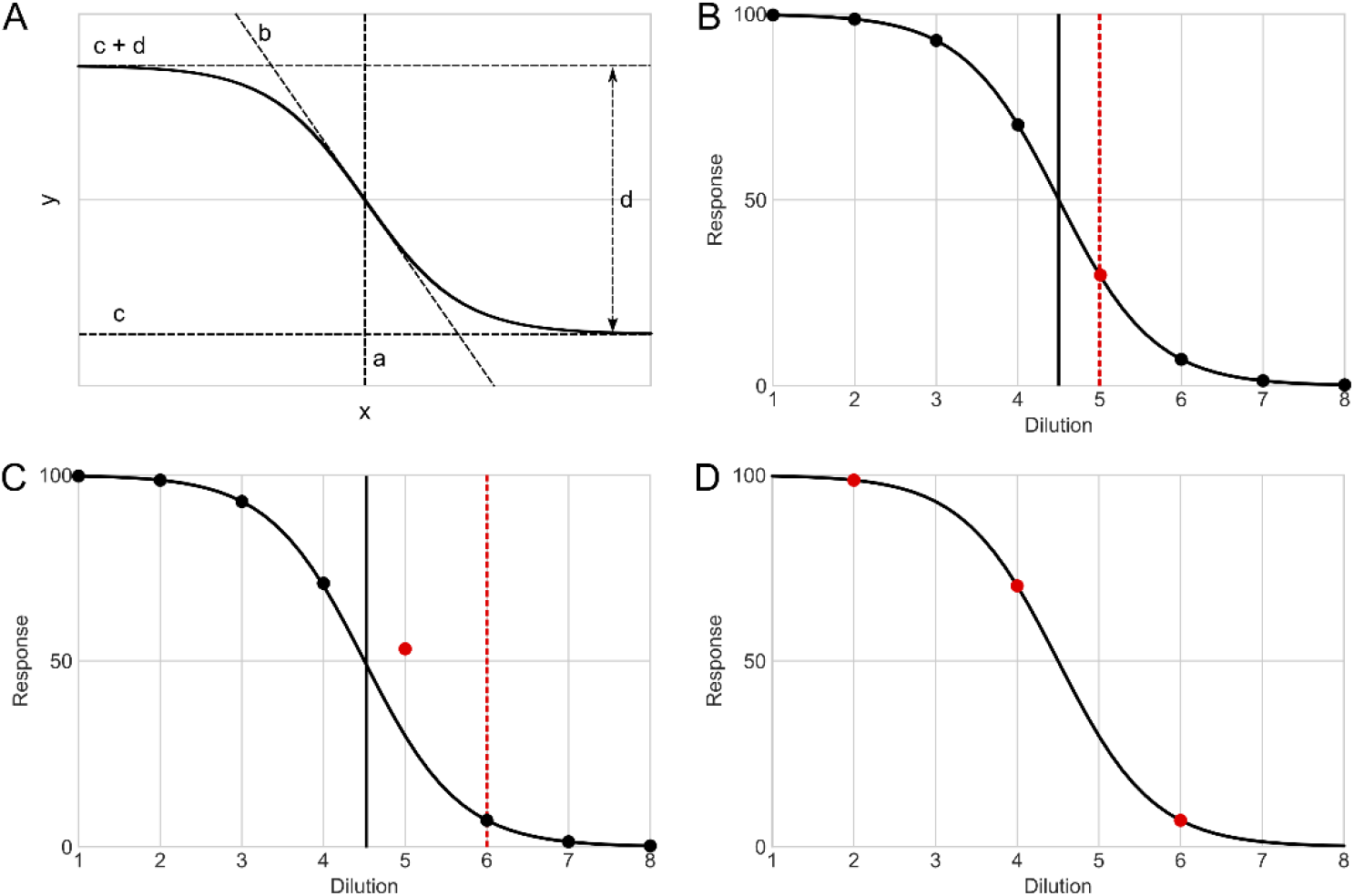
**(A) Four parameter sigmoid function**. y = c+d/(1+exp-b(x-a)). a defines the horizontal location of the curve on the x axis. b sets the gradient of the curve. c sets the minimum y value, and d sets the distance between the minimum and maximum y value. The x-axis shows the index of the dilution series. **(B) Discretizing reduces precision**. The first dilution underneath a 50% response (red point) is 5, but the true inflection point is 4.5 (black vertical line). **(C) Overreliance on individual measurements**. The measurement at dilution 5 is erroneous, so the first dilution underneath a 50% response is 6 (red dotted line), but the true EC50 is 4.5 (black line). **(D) Sigmoid curves can be fit with fewer dilutions than are commonly conducted**. Statistics of interest (e.g. inflection points or endpoints) can then be computed from the curve. Hierarchical modelling of multiple samples that share curve characteristics improves the inference of individual samples.

**Fig. S2.**
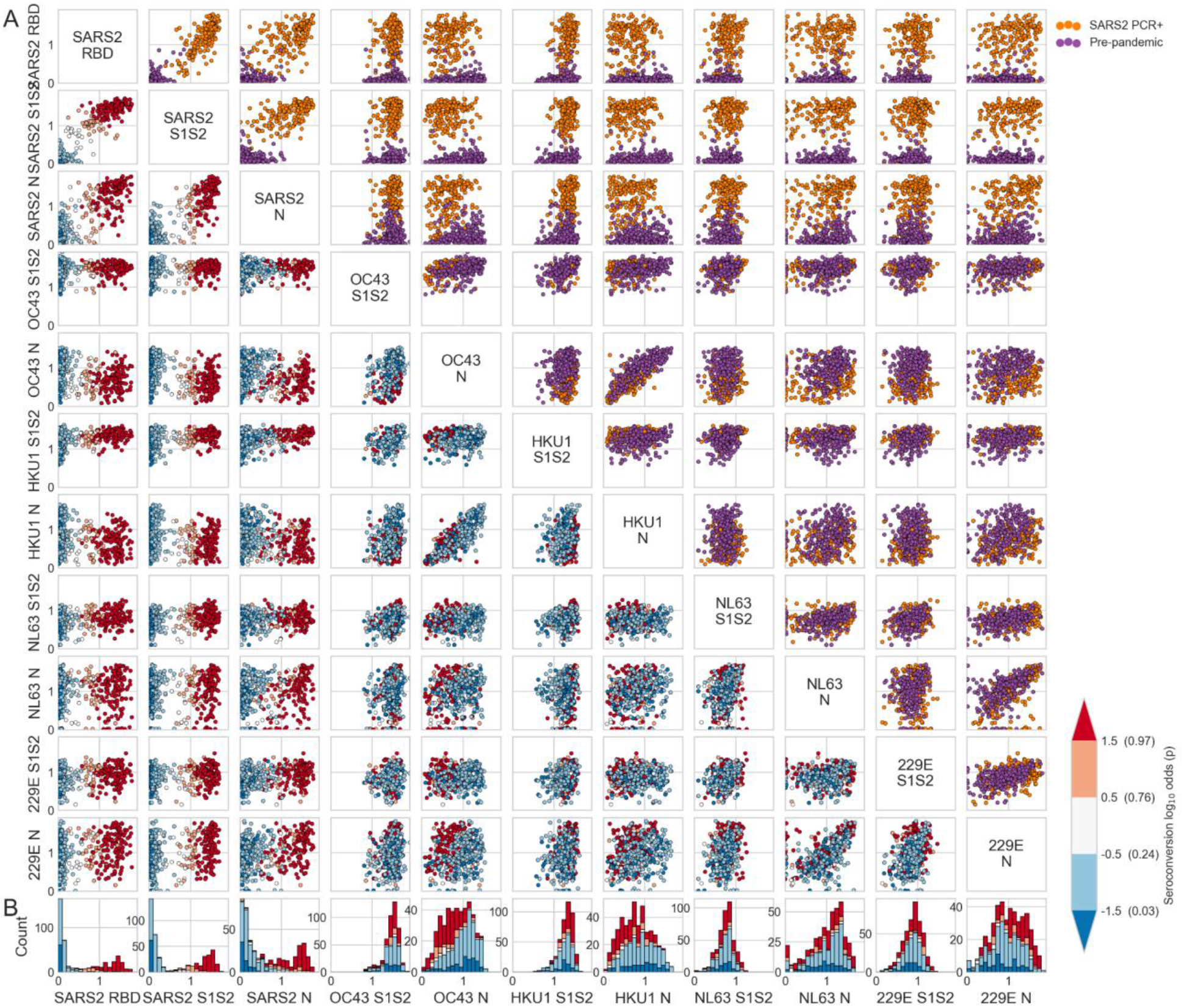
Reactivity to SARS-CoV-2, OC43, HKU1, NL63, 229E spike and N antigens in pre-pandemic and SARS-CoV-2 PCR-positive human serum samples. **(A)** Scatter-matrix of ELISA OD measurements for 1:40 diluted serum samples. Above the main diagonal colors indicate whether a sample was collected before the pandemic or from a SARS-CoV-2 PCR-positive individual. Below the main diagonal, colors indicate SARS-CoV-2 seroconversion log odds. **(B)** Histograms display univariate distributions in each column and are colored by seroconversion log odds.

**Fig. S3.**
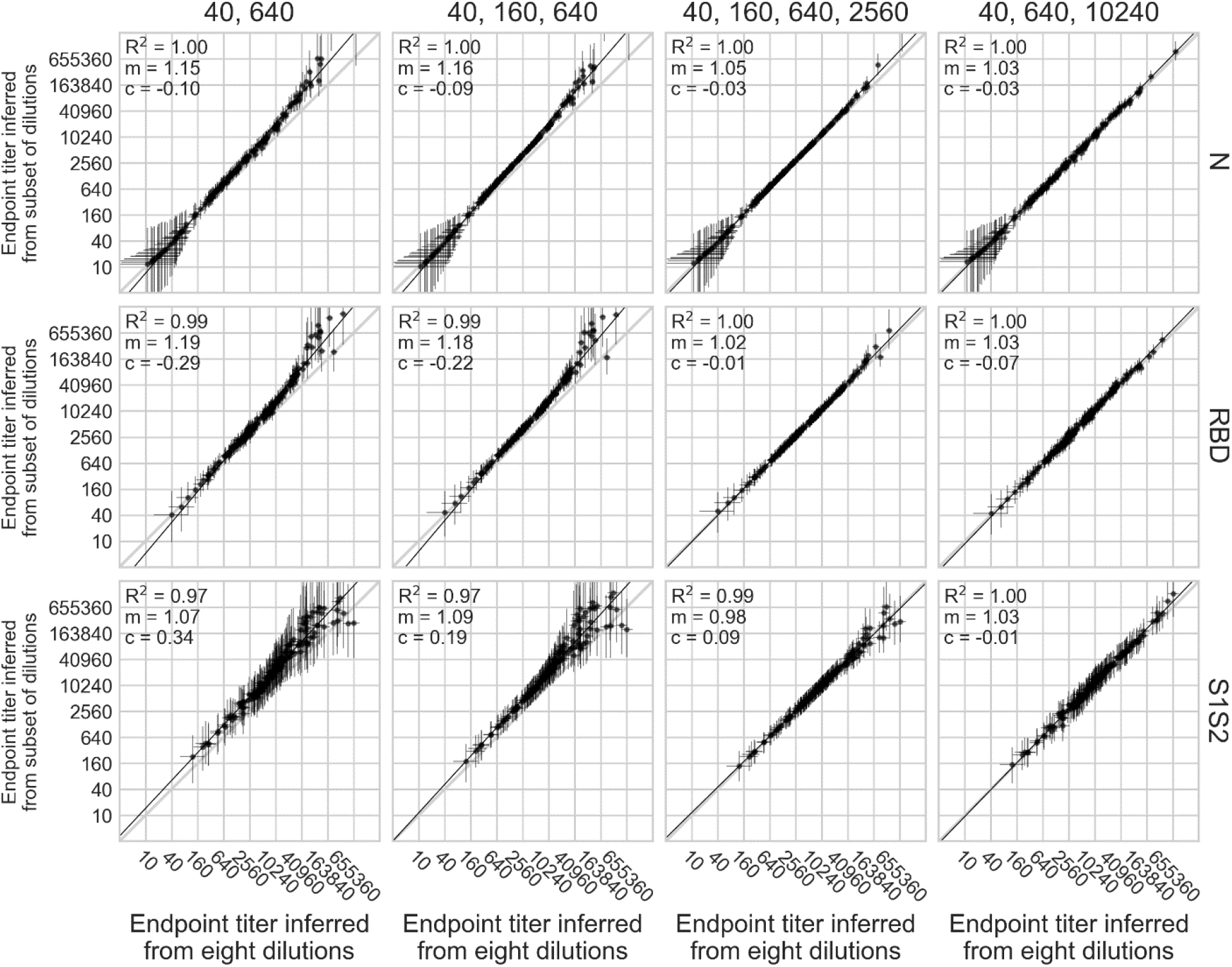
Inferring endpoint titers from minimal dilution series. As Fig. 3, except here we performed the analysis for an arbitrarily chosen endpoint titer, defined here as the serum concentration that would yield an OD reading of 0.1.

**Fig. S4.**
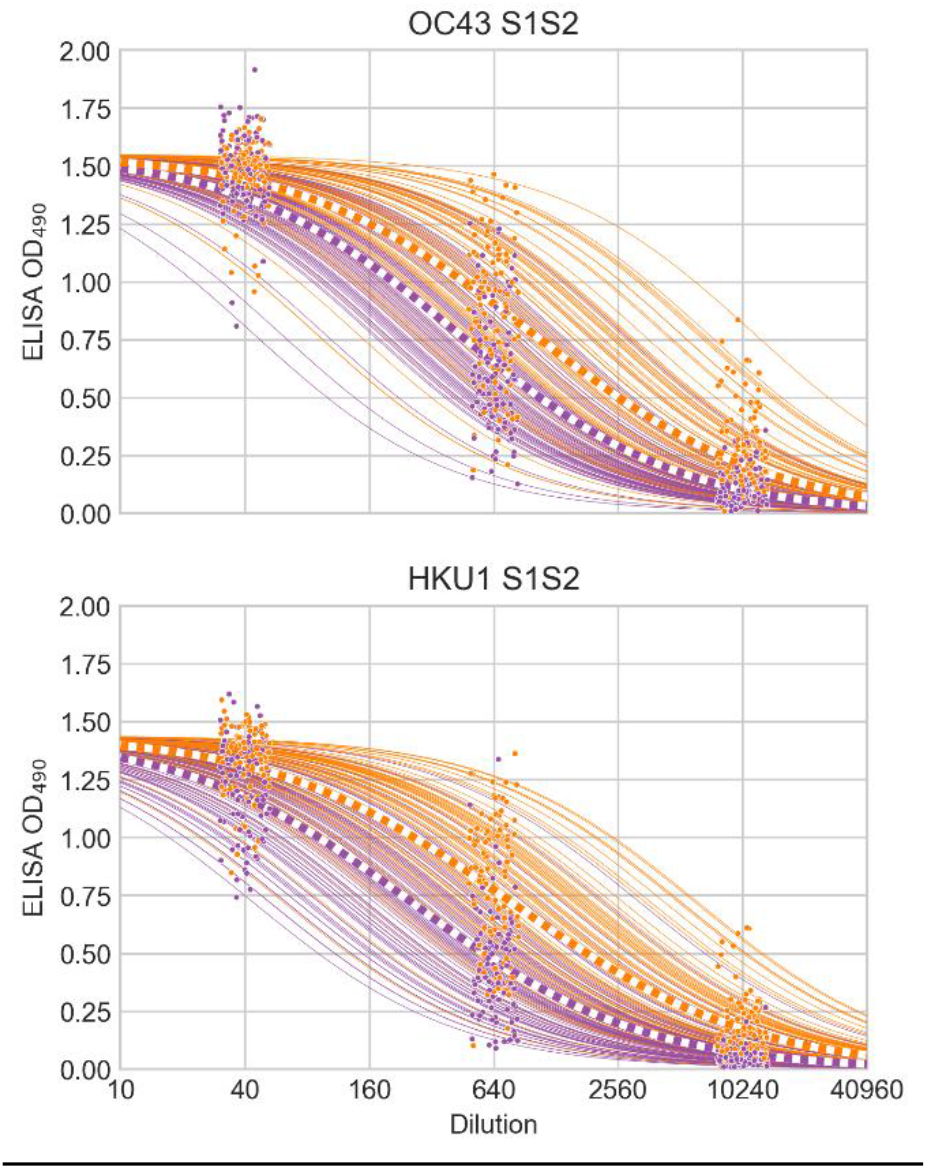
Dilution series of OC43 and HKU1 S1S2. Thin solid lines show sigmoid curves for pre-pandemic (purple) and PCR-positive (orange) samples fit to OD values from their dilution series (points). Thick dashed lines indicate the mean curve of each group. A small amount of x-axis jitter has been added to the points to prevent overplotting. Curves are drawn using the mean of the parameter posterior distributions. The difference between groups for OC43 S1S2 when only looking at OD values from the 1:40 dilution (Fig. 2) is substantially smaller than the difference between the inflection titers (Fig. 5). This is caused by the OD measurement becoming saturated at 1:40. Similarly, for HKU1 the difference between groups is smaller at the single dilution of 1:40 (Fig. 2) than it is when comparing inflection titers (Fig. 5).

**Table S1.** Summary of human serum panels.

| Panel | Description | n |
| --- | --- | --- |
| Pre-pandemic | Obtained before emergence of SARS-CoV-2, between 2013 and April 2019 by the Institute for Influenza Research, UW Madison. | 248 |
| UWH PCR positive | PCR-positive for SARS-CoV-2 from University of Wisconsin Hospital. | 188 |
| Commercial positive | SARS-CoV-2 positive antisera purchased from Lampire Biological Laboratories. | 12 |
| EGH early infection | Paired samples collected from SARS-CoV-2 patients soon after diagnosis at the Eiju General Hospital. | 44 |
| EGH late infection | Paired samples collected from SARS-CoV-2 patients late during infection at the Eiju General Hospital. | 44 |
| MCRI | Samples collected after the emergence of SARS-CoV-2 from participants of a community cohort study by the Marshfield Clinic Research Institute. | 162 |

